# Relationship Between Blood Group and Risk of Infection and Death in COVID-19: a live Meta-Analysis

**DOI:** 10.1101/2020.06.07.20124610

**Authors:** Fateme Pourali, Mahdi Afshari, Reza Alizadeh-Navaei, Javad Javidnia, Mahmood Moosazadeh, Amirhossein Hessami

**Affiliations:** Student Research Committee, School of Medicine, Mazandaran University of Medical Sciences, Sari, Iran; Department of Community Medicine, School of Medicine, Zabol University of Medical Sciences, Zabol, Iran; Gastrointestinal Cancer Research Center, Mazandaran University of Medical Sciences, Sari, Iran; Health Sciences Research center, Addiction Institute, Mazandaran University of Medical Sciences, Sari, Iran; Systematic Review and Meta-Analysis Expert Group (SRMEG), Universal Scientific Education and Research Network (USERN), Tehran, Iran

**Keywords:** COVID-19, Coronavirus, Blood group, Death

## Abstract

**Introduction:** The relationship between ABO blood group and the incidence of COVID-19 infection and death has been investigated in several studies. The reported results were controversial, so the objective of the present study is to assess the relationship between different blood groups and the onset and mortality of COVID-19 infection using meta-analysis method.

**Methods:** We searched the databases using appropriate MeSH terms. We screened articles on the basis of titles, abstracts, and full texts and the articles that met the inclusion criteria were selected. Quality assessment was done with the Newcastle-Ottawa Scale checklist. The estimated frequency of COVID-19 infection and death in terms of ABO blood group and the overall estimate of the odd ratio between blood group with COVID-19 infection and death was done with 95% confidence interval.

**Results:** The pooled frequency of blood groups A, B, O, and AB among COVID-19 infected individuals was estimated as 36.22%, 24.99%, 29.67%, and 9.29% respectively. The frequency of blood groups A, B, O, and AB among the dead cases due to COVID-19 infection was estimated as 40%, 23%, 29%, and 8% respectively. The odd ratio of COVID-19 infection for blood group A versus the other blood groups was estimated 1.16 (CI 95%: 1.02-1.33). The corresponding figures for blood groups O and AB versus other blood groups were estimated as 0.73 (CI 95%: 0.60-0.88) and 1.25(CI 95%: 0.84-1.86) respectively.

**Conclusion:** This meta-analysis showed that individuals with blood group A are at higher risk for COVID-19 infection while those with blood group O are at lower risk. Although the odds ratio of death for AB blood group was non-significant, it was considerable.

## Introduction

The coronaviruses (CoVs) are enveloped, single-stranded, positive-sense RNA viruses that have club-shaped spikes on their surface (1, 2). Before 2002, they were known as one of the causes of the common cold one of their families named *betacoronavirus*, was responsible for more severe issues. In 2002, severe acute respiratory syndrome (SARS), and in 2012, Middle East Respiratory Syndrome (MERS) and recently coronavirus disease 2019(COVID-19) were caused by the same family (1, 3, 4). The virus that causes COVID-19 is named as SARS-CoV-2 or 2019-nCoV (5, 6). World Health Organization (WHO) declared COVID-19 pandemic on 11th of March 2020.According to WHO statistics over 3 million confirmed cases and over 200 thousand deaths were reported by May 6, 2020 (3, 7).

Different risk factors have been reported for COVID-19 morbidity and mortality. These include male, aged over 65 and smoking, and underlying diseases such as hypertension, diabetes, cardiovascular and respiratory diseases (8). Recently some studies found a relationship between the ABO blood group and COVID-19 morbidity and mortality(9-12). ABO blood group have been also reported to be related to different diseases and syndromes such as Norwalk virus, Helicobacter pylori infections, SARS infection (13), acute respiratory distress syndrome(ARDS) (14), influenza (15, 16), acute hypoxemic respiratory failure (17), prostate and bladder cancer (18), gastric cancer and cardiovascular diseases (19).

As mentioned, there are articles emphasizing the hypothesis of a relationship between ABO blood group and COVID-19. However, the results of these articles are not consistent.

Meta-analysis is one of the study designs that combines the results of the preliminary studies and determine a valid estimate. Therefore, our objective is to perform a rapid systematic review and meta-analysis to discover any association between ABO blood group and COVID-19 morbidity and mortality. Note that there are limited primary evidences regarding the association between blood groups and COVID19 infection, upcoming relevant studies will be added to the results of the present meta-analysis.

## Methods

### Search Strategy

A systematic search was carried out in the available databases including PubMed, Scopus, Cochrane library, Web of Science, and also unpublished results in medRxiv. We used all MeSH terms and relevant keywords (COVID19, SARS-CoV-2 infection, COVID-19 virus disease, 2019-nCoV infection, ABO Blood Group System, ABO Factor, Blood Groups, Antigens, Blood Group). All case-control, cohort, and cross-sectional studies until 21^st^ April 2020 were included.

### Criteria for study selection

Inclusion criteria: 1)Studies that reported a relationship between ABO blood group and death due to COVID-19, 2) Studies that reported a relationship between ABO blood group and COVID-19 infection, 3) Studies that reported frequency of COVID-19 among different ABO blood groups and 4)Studies that reported death among COVID-19 infected people.

Exclusion criteria: case-reports and letters to the editors were excluded.

### Data extraction

Data extracted from the primary studies included first author’s name, year of publication, place of the study conduction, type of the study, sampling method, number of participants, number of COVID-19 infection, and death in each blood group, A, B, O, and AB. The required data was entered into Excel spreadsheets.

### Quality assessment

Quality assessment was performed using The Newcastle-Ottawa Scale (NOS). This checklist has three parts: selection, comparability, and exposure. The checklist scores are between 0-9. Studies with a score lower than 5 were excluded. The selection criteria for Selection was 4 in maximum, 2 for Comparability, and 3 for Exposure (20). Quality assessment was performed by two authors independently.

### Data analysis

Data analysis was performed using Stata ver.11. Heterogeneity between studies was assessed using Cochrane’s Q test and I-squared (I^2^) test. Standard error was calculated to assess the frequency of COVID-19 infection and death in each blood group. The overall estimate of the frequency of COVID-19 infection and death with 95% confidence interval in each ABO blood group was calculated, using random effect model. To assess the relationship between COVID-19 infection and mortality with blood group, the required data was extracted in binary tables. Using the *Metan* command, the random effect model, and reverse variance, the point odd ratio and 95% confidence interval was illustrated on forest plots. In these plots, the size of the square shows the weight of each study and the lines beside it show the 95% confidence interval. In the cases that confidence interval didn’t include number 1, the difference was considered statistically significant.

## Results

By searching in databases, 318 studies were found. After removing duplicates, checking the titles, abstracts and full texts of the remaining articles, 314 irrelevant or duplicate papers were excluded. Quality assessment scores for the four final selected studies in the meta-analysis were above 5. The eligible studies included two case-controls, one cohort and one cross sectional studies. Of them, (figure 1). The studies were done in China (3 studies) or the U.S.A (one study). In total, 7 data were found, from hospitals in Wuhan, Shenzhen, Xi’an, Beijing, and New York City. Totally,139,128 participants were enrolled in these studies 135,940 of which were controls.

**Figure 1.**
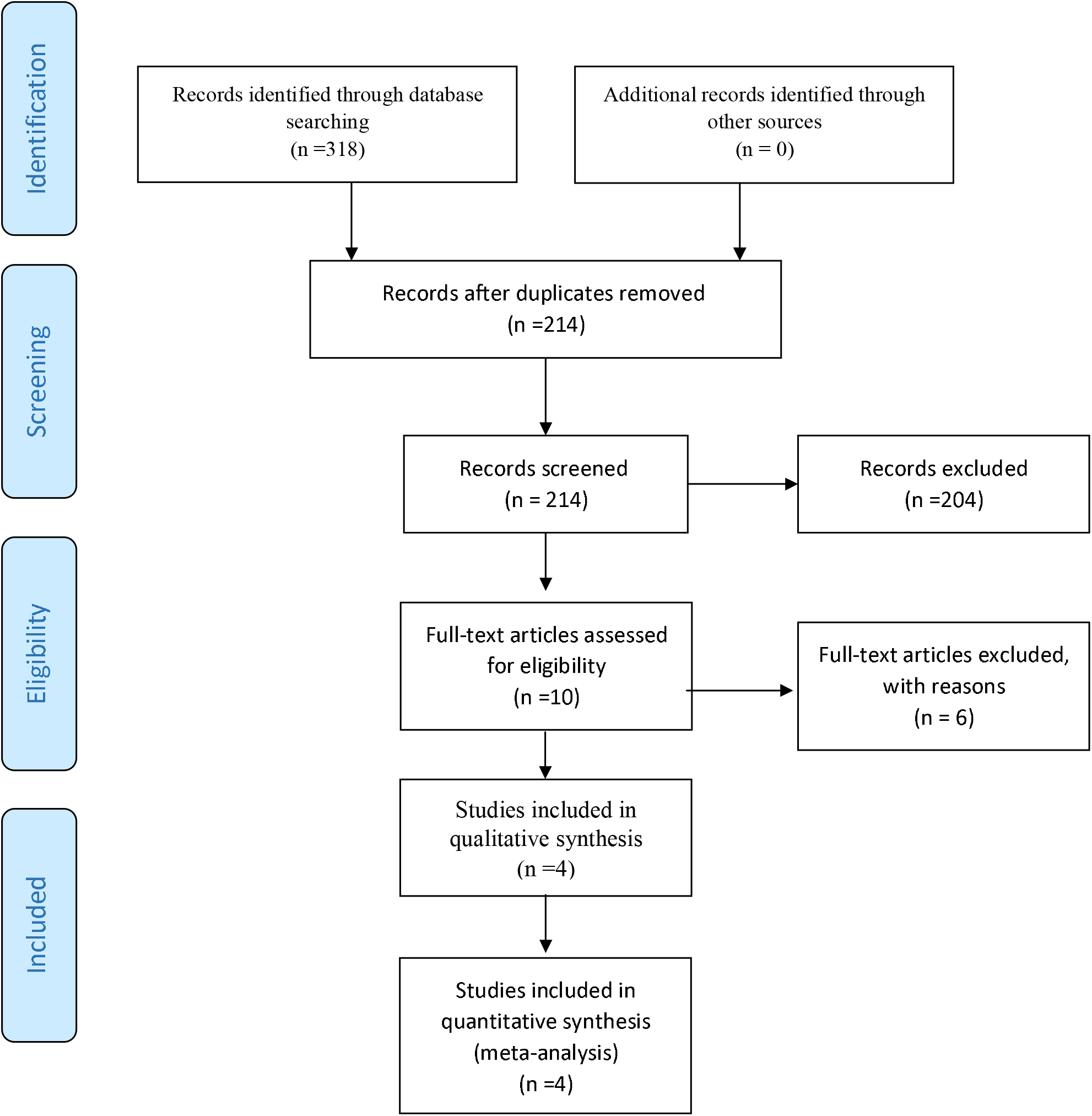
Flowchart of study selection

In 7 evidence included in this meta-analysis, the frequency of blood group A among COVID-19 infected people had been reported between 28.77% and 44.44%. Combining these results using random effect model (I-square:41.5%, Q:10.25, P-value:0.115), the pooled frequency of blood group A among COVID-19 infected people, was estimated as 36.22% (95% CI:32.81,39.63) (figure 2-A).

**Figure 2.**
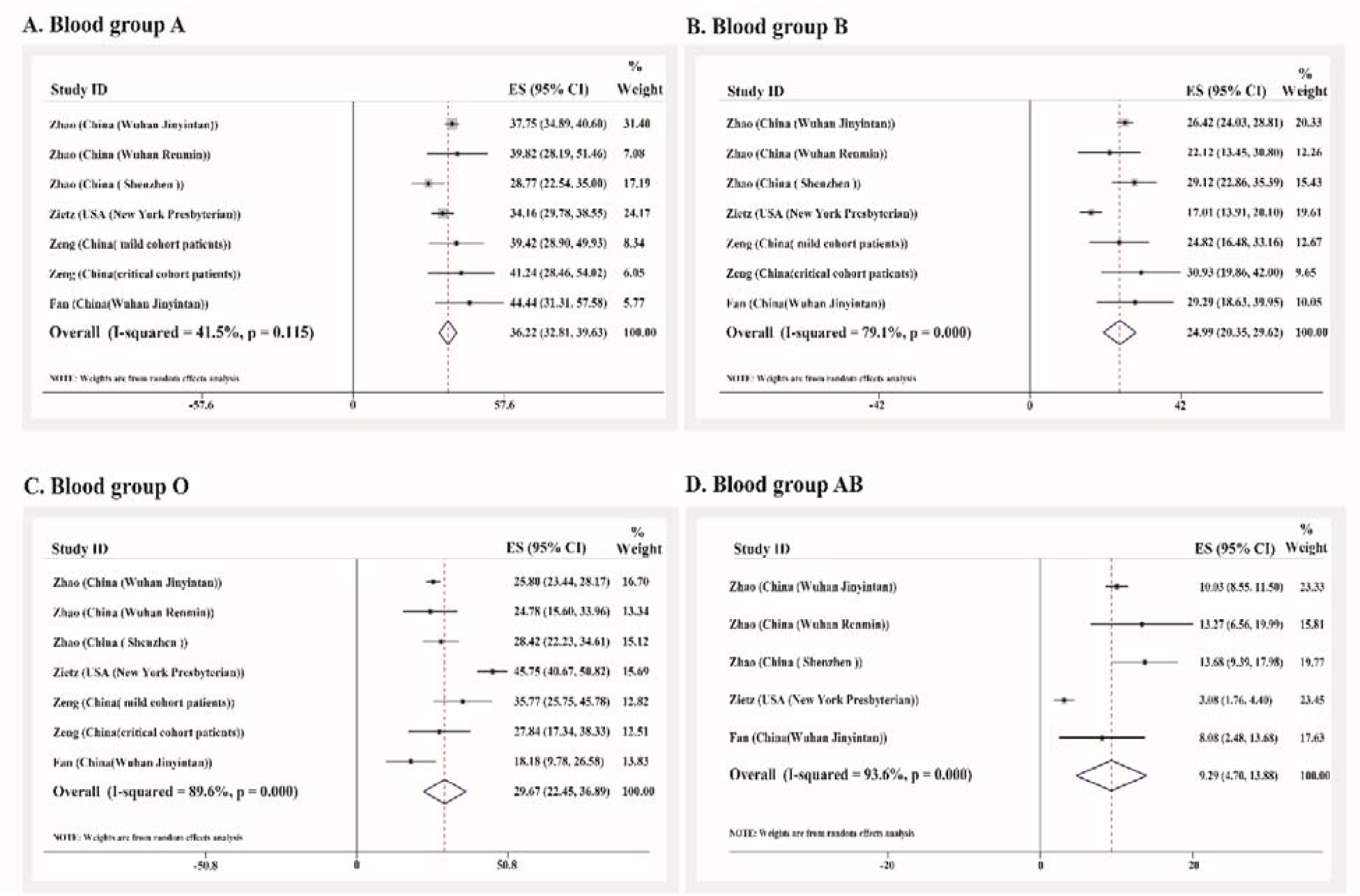
forest plot of blood group A, B, O, and AB frequency with 95% confidence interval among COVID-19 infected individuals, in each evidence and overall

The frequency of blood group B among COVID-19 infected people had been reported in the seven evidences between 17.01% and 30.93%. Combining these results with random effect model (I-squared:79.1%, Q:28.70, P-value<0.001) the total frequency of blood group B among all COVID-19 infected people, was estimated as 24.99% (95% CI:20.35, 29.62) (figure 2-B).

In 7 evidence included in this meta-analysis, the frequency of blood group O among COVID-19 infected people had been reported from 18.18% to 45.75%. Combining these results with random effect model (I-squared:89.6%, Q:57.77, P-value<0.001), the pooled frequency of blood group O among COVID-19 infected people, was estimated as 29.67% (95% CI:22.45, 36.89) (figure 2-C).

Of the evidence included in this meta-analysis, five studies had reported the frequency of blood group AB among COVID-19 infected people varied between 3.08% and 13.68%. Combining the results with random effect model (I-squared:93.6%, Q:62.72, P-value<0.001), the frequency of blood group AB among COVID-19 infected people, was estimated as 9.29% (95% CI:4.70, 13.88) (figure 2-D).

The odds for COVID-19 infection among blood group A versus non-A blood groups was extracted from four evidences one of which was statistically significant. Combining the primary odds ratios using random effect model (I-square=47.1%, Q=5.67, P=0.129), the pooled odd ratio for blood group A was estimated as 1.16 (95% CI: 1.02-1.33) (figure 3-A).

**Figure 3.**
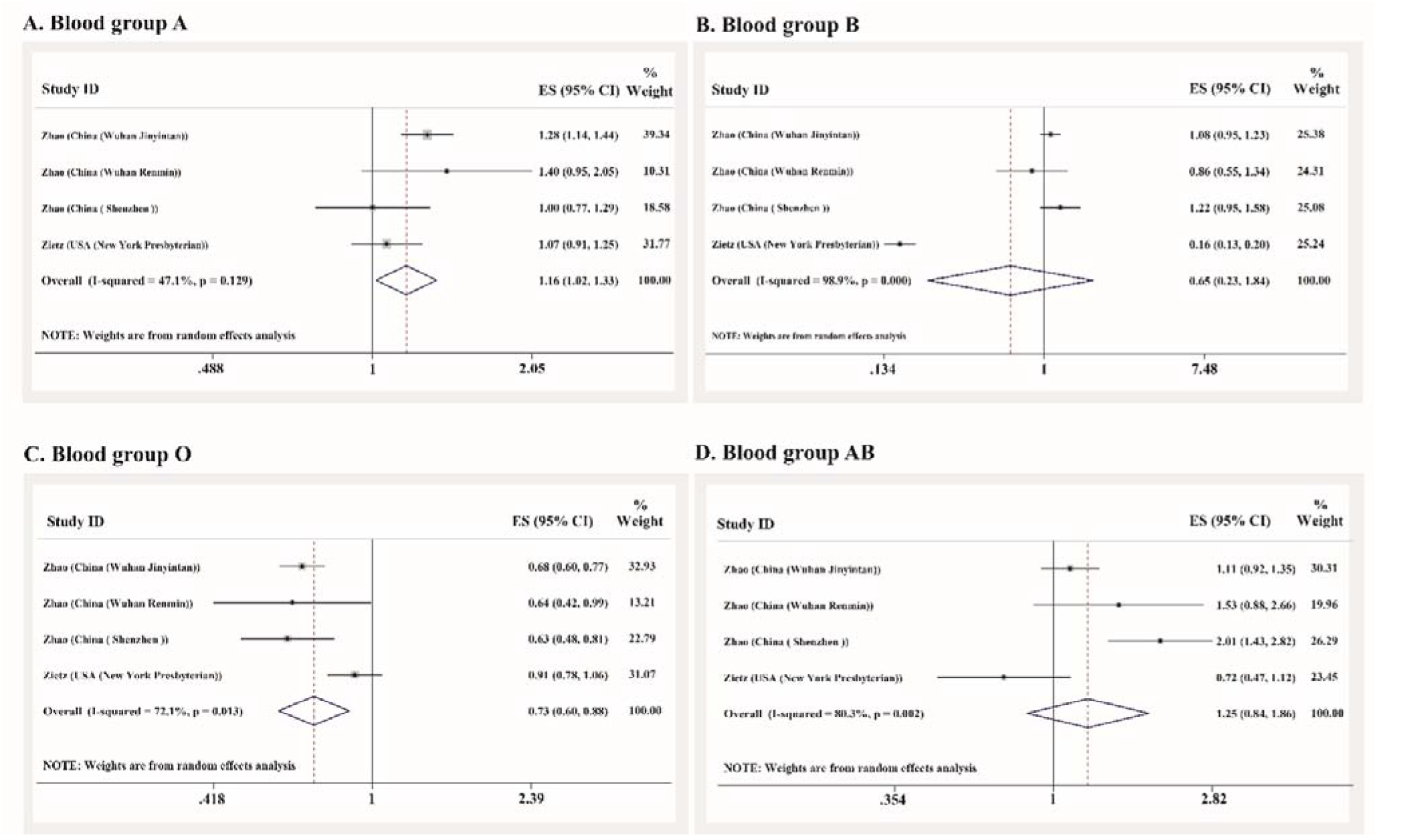
The point and pooled odd ratios for COVID-19 infection among individuals with blood groups A, B, O, and AB

The odds of COVID-19 infection among blood group B versus non-B blood groups was extracted from four evidences. it was lower in B blood group than non-B blood groups in two studies, one of which was statistically significant. Combining these results with random effect model (I-square=98.9%, Q=266.1, P<0.001), the odd ratio for blood group B was estimated as 0.65 (95% CI: 0.23-1.84) (figure 3-B).

The odds of COVID-19 infection among blood group O versus non-O blood groups had been reported in four evidence all of which reported lower odds of COVID-19 infection among subjects with blood group O. Three of these associations were statistically significant. By combining these results with random effect model (I-square=72.1%, Q=10.76, P=0.013), the odd ratio for blood group O was estimated as 0.73 (95% CI: 0.60-0.88) (figure 3-C).

The odds of COVID-19 infection among patients with and without blood group AB had been reported in four evidences just one of which was statistically significant. Combining these results using random effect model (I-square=80.3%, Q=15.22, P=0.002), the odd ratio for having blood group AB was estimated as 1.25 (95% CI: 0.84-1.86) (figure 3-D).

The odds for death among COVID-19 infected people with blood group A versus non-A blood group had been reported by two evidences one of which reported higher chance of mortality among patients with blood group A compare to those without. However, it was not statistically significant. Combining the results of these two evidences, applying random effect model (I-square=0%, Q=0.41, P=0.522), the odd ratio for death among COVID-19 infected people having blood group A was estimated as 1.12 (95% CI: 0.87, 1.45).

Just two studies had compared patients with and without blood group B in term of the odds of COVID-19 infection both of them showed lower odds of death among people with blood group B. but the results were not statistically significant. Combining the results of these two evidences using random effect model (I-square=0%, Q=0.01, P=0.914), the odd ratio for blood group B versus was estimated as 0.87 (95% CI: 0.65, 1.18)

The odds ratio for death of COVID-19 infected people following blood group O had been reported by two evidences, both of them showed negative association. However, the results were not statistically significant. Combining the results of these two evidences, applying random effect model (I-square=0%, Q=0.00, P=0.998), the odd ratio for blood group O was estimated as 0.97 (95% CI: 0.74, 1.27)

The odds ratios for death among COVID-19 infected people following blood group AB had been reported by two evidences. Both of these studies reported more chance of developing death among patients with AB group. However, they were not statistically significant. Combining the results of these two evidences, applying random effect model (I-square=65.5%, Q=2.90, P=0.088), the odds ratio for blood group AB was estimated as 1.33 (95% CI: 0.51, 3.46) (table 1).

**Table 1.** The overall and point odds ratios with 95% confidence intervals of death due to COVID-19 infection in

| Blood groups | Number of evidences | OR | 95% CI | Heterogeneity index |  |  |
| --- | --- | --- | --- | --- | --- | --- |
|  |  |  |  | I-square (%) | Q | P-value |
| A (non-A) | 2 | 1.12 | 0.87-1.45 | 0 | 0.41 | 0.522 |
| B (non-B) | 2 | 0.87 | 0.65-1.18 | 0 | 0.01 | 0.914 |
| O (non-O) | 2 | 0.97 | 0.74-1.27 | 0 | 0.00 | 0.998 |
| AB (non-AB) | 2 | 1.33 | 0.51-3.46 | 65.5 | 2.90 | 0.088 |

terms of blood groups

In three studies, the frequency of death among individuals with COVID-19 infection was reported in terms of different blood groups. Combining the results of these articles, the frequency of death due to COVID-19 among patients with A, B, O and AB groups were estimated as 40% (95% CI : 35-46%), 23% (95% CI:15-30%), O 29% (95% CI: 16-42%), and 8% (95% CI:5-11%) respectively.

## Discussion

In this study, we found that blood group A was a partial risk factor for COVID-19 infection (OR=1.16(1.02-1.33)) while blood group O was a protective factor (OR=0.73(0.60-0.88)). Moreover, B and AB blood groups were not significantly associated with COVID-19 infection.

In a letter to the editor investigating ABO blood groups and susceptibility to SARS in 2005, 45 hospital staff in contact with a patient without any protective clothes were checked. They were tested for SARS-CoV IgG antibody(13). In our included studies, cases were tested with molecular methods or clinical diagnostic criteria(6, 9, 10, 12) The results for this study showed that individuals with blood group O, were less susceptible to SARS infection(13), which is consistent with our results on the cases with SARS-CoV-2 infection. In that study, the results were not statistically significant for blood group B and were undefined for blood group A and AB (13).

According to the results of a commentary, blood group A was considered to have more attachment molecules on the vascular wall so the adhesion and inflammation is increased and the symptoms of COVID-19 would be more severe than non-A blood groups. This study suggests blood group A as a predisposing factor not a risk factor (21).

Angiotensin converting enzyme 2 (ACE2) has been reported as the SARS-CoV receptor. And the receptor binding domain (RBD) is presented on the S proteins of the coronaviruses. If a factor could prevent interaction between the RBD of the virus and its receptor on the host cells, it could prevent infection. Blood group antigens are expressed on almost all cells including epithelial cells which are the host cells for SARS-CoV. So during the replication, these carbohydrates could express on S proteins and the natural antibodies in blood groups O, B, A could bind to them and prevent RBD interaction with ACE2 leading to infection prevent (22).

As SARS-CoV and SARS-CoV-2 are from the same family (betacoronavirus) (4) and have similarities in the structure of RBD, ACE2 has been also suggested as the receptor for SRAS-CoV-2 (6). Therefore, the same mechanism might explain the lower susceptibility of blood group O to SARS-CoV-2.

In explaining the higher risk for blood group A, lack of these antibodies can be expected, although it needs further studies to be confirmed (9).

Except for binding to ACE2, the virus can bind to the ABO carbohydrates. Lack of these antigens on the blood group O leads to the least molecular contact with the virus and lower risk of getting infection (23, 24).

individuals with blood group O have a lower ACE level. ACE is an enzyme that activates angiotensin. Thus, the lower level of this enzyme can reduce the risk of hypertension which is a COVID-19 risk factor. Although ACE2 is the virus receptor, it can have some benefits. For example it can attenuate inflammatory response and redox stress and counter balance ACE effect and in the case of lower ACE level, it can work even more effectively (8, 21). In addition, individuals with blood group O, have a higher interleukin 6 (IL-6) (25). IL-6 is a proinflammatory cytokine that can be produced by many cells and has an important role in cell defense in acute phase (26).

It should be noted that all of these mechanisms should be more investigated to be accepted.

Among COVID-19 infected individuals and the dead cases, blood groups A and AB had the highest and lowest frequencies respectively. But blood group AB wasn’t significantly associated with the risk of COVID-19. One explanation for this controversy is that blood group AB is less frequent in the society.

The mortality rate of COVID-19 was reported 11% in a study carried out on 99 cases in china (27).

COVID-19 mortality is probably higher in older people, obese individuals and those who have underlying diseases(27). No statistically significant relationship was found between the blood group and mortality of COVID-19 in our study.

Although we didn’t find a statistically significant relationship between blood group A and COVID-19 mortality, one study suggests higher mortality among blood group A individuals. It might be because of failure of pulmonary microcirculation and the higher risk of thrombosis in blood group A, as microthrombi have been seen in lungs and kidneys(28)

Higher total mortality due to medical causes among non-O blood groups has been reported. This specially includes more cardiovascular mortality which can be because of the higher levels of von Willebrand (vWF) and VIII factors in these individuals. Cancer mortality has also been reported to be higher among non-O blood groups specially blood group. However, these observed associations were not statistically significant (18, 19).

Further studies are needed to understand the relationship between blood group and COVID-19 mortality.

## Limitation and suggestion

The primary studies entered into the present meta-analysis have not been peer-reviewed yet.

Confounding factors: diabetes, hypertension, cardiovascular diseases etc. were not adjusted in the study, because of incomplete information.

Another limitation is that blood group O is more frequent than the other blood groups.

More observational studies with large-scale samples and considering the confounding factors are needed to prove the relationship. In vitro studies are also needed to understand the exact mechanism of protective or deteriorating effect of blood groups.

## Data Availability

Data are available upon request from the corresponding author.

## References

1. Malik Y. Properties of Coronavirus and SARS-CoV-2. The Malaysian Journal of Pathology. 2020;42(1):3–11.

2. Pal M, Berhanu G, Desalegn C, Kandi V. Severe Acute Respiratory Syndrome Coronavirus-2 (SARS-CoV-2): An Update. Cureus. 2020;12(3).

3. Baloch S, Baloch MA, Zheng T, Pei X. The Coronavirus Disease 2019 (COVID-19) Pandemic. The Tohoku Journal of Experimental Medicine. 2020;250(4):271–8.

4. Zainol Rashid Z, Othman SN, Abdul Samat MN, Ali UK, Wong KK. Diagnostic performance of COVID-19 serology assays. Malays J Pathol. 2020;42(1):13–21.

5. Leung C. Risk factors for predicting mortality in elderly patients with COVID-19: a review of clinical data in China. Mechanisms of Ageing and Development. 2020:111255.

6. Lu R, Zhao X, Li J, Niu P, Yang B, Wu H, et al. Genomic characterisation and epidemiology of 2019 novel coronavirus: implications for virus origins and receptor binding. The Lancet. 2020;395(10224):565–74.

7. WHO.int.Coronavirus disease (COVID-19) Situation dashboard 2020 [Available from: https://covid19.who.int/.

8. Zheng Z, Peng F, Xu B, Zhao J, Liu H, Peng J, et al. Risk factors of critical & mortal COVID-19 cases: A systematic literature review and meta-analysis. Journal of Infection. 2020.

9. Chen J, Fan H, Zhang L, Huang B, Zhu M, Zhou Y, et al. Retrospective Analysis of Clinical Features in 101 Death Cases with COVID-19. medRxiv. 2020.

10. Zeng X, Fan H, Lu D, Huang F, Meng X, Li Z, et al. Association between ABO blood groups and clinical outcome of coronavirus disease 2019: Evidence from two cohorts. medRxiv. 2020.

11. Zhao J, Yang Y, Huang H-P, Li D, Gu D-F, Lu X-F, et al. Relationship between the ABO Blood Group and the COVID-19 Susceptibility. medRxiv. 2020.

12. Zietz M, Tatonetti NP. Testing the association between blood type and COVID-19 infection, intubation, and death. medRxiv. 2020.

13. Cheng Y, Cheng G, Chui C, Lau F, Chan PK, Ng MH, et al. ABO blood group and susceptibility to severe acute respiratory syndrome. Jama. 2005;293(12):1447–51.

14. Reilly J, Meyer N, Shashaty M, Feng R, Lanken P, Gallop R, et al. ABO blood type A is associated with increased risk of acute respiratory distress syndrome in caucasians following both major trauma and severe sepsis. Chest. 2014;145(4):753–61.

15. Lebiush M, Rannon L, Kark J. The relationship between epidemic influenza A (H 1 N 1) and ABO blood groups. Epidemiology & Infection. 1981;87(1):139–46.

16. Mackenzie J, Fimmel P. The effect of ABO blood groups on the incidence of epidemic influenza and on the response to live attenuated and detergent split influenza virus vaccines. Epidemiology & Infection. 1978;80(1):21–30.

17. Rezoagli E, Gatti S, Villa S, Villa G, Muttini S, Rossi F, et al. ABO blood types and major outcomes in patients with acute hypoxaemic respiratory failure: A multicenter retrospective cohort study. PloS one. 2018;13(10).

18. Stakišaitis D, Juknevičienė M, Ulys A, Žaliūnienė D, Stanislovaitienė D, Šepetienė R, et al. ABO blood group polymorphism has an impact on prostate, kidney and bladder cancer in association with longevity. Oncology letters. 2018;16(1):1321–31.

19. Etemadi A, Kamangar F, Islami F, Poustchi H, Pourshams A, Brennan P, et al. Mortality and cancer in relation to ABO blood group phenotypes in the Golestan Cohort Study. BMC Med. 2015;13:8-.

20. Stang A. Critical evaluation of the Newcastle-Ottawa scale for the assessment of the quality of nonrandomized studies in meta-analyses. European journal of epidemiology. 2010;25(9):603–5.

21. Dai X. ABO blood group predisposes to COVID-19 severity and cardiovascular diseases. European Journal of Preventive Cardiology. 2020:2047487320922370.

22. Guillon P, Clément M, Sébille V, Rivain J-G, Chou C-F, Ruvoën-Clouet N, et al. Inhibition of the interaction between the SARS-CoV spike protein and its cellular receptor by anti-histo-blood group antibodies. Glycobiology. 2008;18(12):1085-93

23. Mattos LCd, Moreira HW. Genetic of the ABO blood system and its link with the immune system. Revista Brasileira de Hematologia e Hemoterapia. 2004;26(1):60-3.

24. P A. How blood group A might be a risk and blood group O be protected from coronavirus (COVID-19) infections (how the virus invades the human body via ABO(H) blood group carbohydrates): figshare; 2020 2020 [Available from: https://figshare.com/articles/How_blood_group_O_could_be_protected_from_Coronavirus_Covid-19_infections/12019035/122.

25. Naitza S, Porcu E, Steri M, Taub DD, Mulas A, Xiao X, et al. A genome-wide association scan on the levels of markers of inflammation in Sardinians reveals associations that underpin its complex regulation. PLoS genetics. 2012;8(1).

26. Zhang C, Wu Z, Li J-W, Zhao H, Wang G-Q. The cytokine release syndrome (CRS) of severe COVID-19 and Interleukin-6 receptor (IL-6R) antagonist Tocilizumab may be the key to reduce the mortality. International Journal of Antimicrobial Agents. 2020:105954.

27. Chen N, Zhou M, Dong X, Qu J, Gong F, Han Y, et al. Epidemiological and clinical characteristics of 99 cases of 2019 novel coronavirus pneumonia in Wuhan, China: a descriptive study. The Lancet. 2020;395(10223):507-13.

28. Menter T, Haslbauer JD, Nienhold R, Savic S, Hopfer H, Deigendesch N, et al. Post-mortem examination of COVID19 patients reveals diffuse alveolar damage with severe capillary congestion and variegated findings of lungs and other organs suggesting vascular dysfunction. Histopathology. 2020.

